# Humoral Immunity Elicited by the XBB.1.5 Monovalent COVID-19 Vaccine

**DOI:** 10.1101/2024.03.25.24304857

**Authors:** Xammy Huu Nguyenla, Timothy A. Bates, Mila Trank-Greene, Mastura Wahedi, Fikadu G. Tafesse, Marcel Curlin

## Abstract

As novel SARS-CoV-2 variants continue to emerge, the updated XBB.1.5 monovalent vaccines remain to be evaluated in terms of immunogenicity against live clinical isolates. We report boosting of IgG(2.1X), IgA(1.5X), and total IgG/A/M(1.7X) antibodies targeting the spike receptor-binding domain and neutralizing titers against WA1(2.2X), XBB.1.5(7.4X), EG.5.1(10.5X), and JN.1(4.7X) variants.

## Introduction

The updated monovalent COVID-19 vaccines containing the XBB.1.5 variant spike protein were recently approved, yet uptake has been hesitant^1^. Clear evaluation of the immunogenicity of variant-adapted vaccines is important to trust in future COVID-19 immunization campaigns, especially with the emergence of neutralization-evading variants like JN.1. Studies have demonstrated induction of antibodies capable of neutralizing variant spike proteins^2–4^; however, such studies utilize pseudo-typed virus that recombinantly express variant spike proteins as opposed to true SARS-CoV-2. We evaluate the immunogenicity of XBB.1.5 vaccination in humans using live clinical isolates of SARS-CoV-2, which more fully captures the biology of virus neutralization.

## Methods

Healthcare workers were recruited at Oregon Health & Science University (OHSU) between October-November 2023. Paired serum samples were collected on the day of XBB.1.5 monovalent vaccine (Moderna) administration and ∼21 days after vaccination. Anti-nucleocapsid antibodies were detected by enzyme-linked immunosorbent assay (ELISA) to identify recent infection. IgG, IgA, IgM, and total IgG/A/M antibody titers against the ancestral spike RBD were determined by 50% ELISA effective concentrations (EC50). Live SARS-CoV-2 neutralizing antibody titers were determined by 50% focus reduction neutralization tests (FRNT50) against ancestral SARS-CoV-2 (WA1) and variants (XBB.1.5, EG.5.1, and JN.1) (**Figure 1A)**. Reported p-values are the results of a restricted effect maximum likelihood model or repeated measures ANOVA with Šídák’s multiple comparisons tests. The OHSU Institutional Review Board approved this study, and written informed consent was obtained from participants. More details on laboratory and analysis methods can be found in the Supplemental Methods.

**Figure 1.**
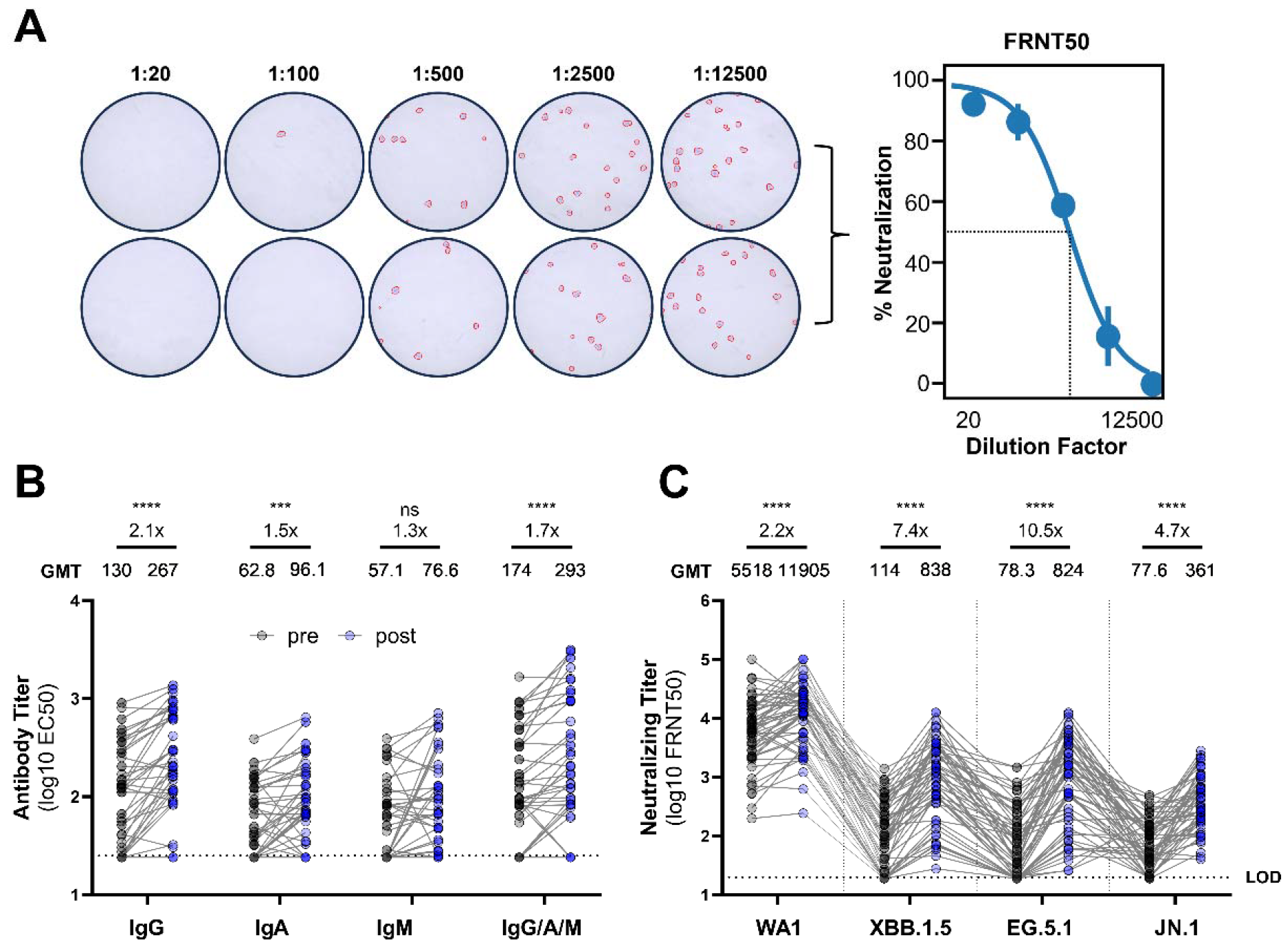
XBB.1.5 monovalent vaccination boosts spike RBD-binding isotypes and neutralizing antibodies against live SARS-CoV-2. (a) Representative focus reduction neutralization test (FRNT) results showing wells infected with live SARS-CoV-2 after incubation with serially diluted serum in duplicate, which were stained and counted to produce the representative FRNT curve. (b) Serum antibody isotype titers against spike RBD were determined by ELISA and reported as EC50. (c) Live SARS-CoV-2 neutralization by serum antibodies was assessed by FRNT and reported as FRNT50s. The dotted lines indicate assay lower limits of detection. Geometric mean titers (GMT) for each bar were calculated in GraphPad Prism. Boost ratios were calculated by dividing the post-XBB.1.5 vaccination GMT (post) by pre-vaccination GMT (pre). Reported p-values are the result of a restricted effect maximum likelihood model (b) or one-way repeated measures ANOVA (c) with Šídák’s multiple comparisons tests (****, p<0.0001; ***, p<0.001; ns=not significant, p>0.05). All individual data points are displayed as filled circles.

## Results

Fifty-five individuals enrolled (mean age, 53 years; 37 [67%] women). 11 (20%) of pre-boost samples and 15 (27%) of post-boost samples were positive for anti-nucleocapsid antibodies. These samples were included in final analysis in order to report generalized boosting in a population with heterogenous exposure history; however, removing these participants from analysis resulted in similar induction of antibodies by XBB.1.5 vaccination (**Supplemental Figure 1A-B**). As expected, the XBB.1.5 vaccine boosted total serum IgG/A/M antibodies targeting the spike RBD [post-boost GMT 293 (95% CI: 195-442) versus pre-boost GMT 174 (124-244); 1.7-fold change; p<0.0001]. IgG isotypes demonstrated a greater increase [post-boost GMT 267 (196-363) versus pre-boost GMT 130 (95.7-176); 2.1-fold change; p<0.0001] than IgA [post-boost GMT 96.1 (74.6-124) versus pre-boost GMT 62.8 (50.3-78.3); 1.5-fold change; p=.0002], possibly due to the intramuscular route of administration as opposed to mucosal vaccination. IgM isotypes trended toward a slight increase [post-boost GMT 76.6 (57.6-102) versus pre-boost GMT 57.1 (44.5-73.2); 1.3-fold change; p=.1548] likely due to their short-lived nature (**Figure 1B**). Importantly, the XBB.1.5 vaccine boosted neutralizing titers against the ancestral WA1 [post-boost GMT 11905 (8454-16766) versus pre-boost GMT 5518 (3899-7809); 2.1-fold change; p<0.0001] and the vaccine-matched XBB.1.5 variant [post-boost GMT 838 (548-1281) versus pre-boost GMT 114 (80.9-162); 7.4 fold-change; p<0.0001]. The vaccine also significantly boosted neutralizing titers against EG.5.1 (post-boost GMT 824 (518-1311) versus pre-boost GMT 78.3 (55.0-112); 10.5 fold-change; p<.0001] and the currently dominant JN.1 (post-boost GMT 361 (270-483) versus pre-boost GMT 77.6 (60.7-99.2); 4.7 fold-change; p<.0001) (**Figure 1C**). To assess changes in the proportion of serum antibodies with neutralizing capacity, the serum neutralizing titer against a given variant was divided by the total IgG/A/M titer to produce a neutralizing potency index (NPI). The NPI against the ancestral WA1 strain was unchanged by XBB.1.5 monovalent vaccination [post-boost GM 40.6 (25.8-63.9) versus pre-boost GM 31.8 (22.0-45.8); p=2931]. This is likely explained by pre-existing neutralizing immunity that is dominated by responses against WA1 epitopes due to prior COVID-19 vaccination with ancestral spike protein and/or historic infections by ancestral SARS-CoV-2 strains. Importantly, the XBB.1.5 vaccine elicits an increase in NPI against XBB.1.5 [post-boost GM 2.86 (1.72-4.76) versus pre-boost GM 0.658 (0.441-0.982); p<0.0001], EG.5.1 [post-boost GM 2.81 (1.63-4.85) versus pre-boost GM 0.451 (0.295-0.689); p<0.0001], and JN.1 [post-boost GM 1.23 (0.781-1.95) versus pre-boost GM 0.447 (0.306-0.652); p<.0001] (**Supplemental Figure 1C**).

## Discussion

Overall, these data provide direct evidence for the immunogenicity of the XBB.1.5 monovalent vaccine against live clinical isolates of SARS-CoV-2. Neutralizing antibodies were boosted against the ancestral WA1 strain, the vaccine-matched XBB.1.5, and emergent EG.5.1 and JN.1, suggesting that updated vaccines may enhance protection against infection by historic and vaccine-matched strains as well as novel, emerging variants. Furthermore, we demonstrate that individuals have low ratios of antibodies capable of neutralizing XBB.1.5, EG.5.1, and JN.1 prior to vaccination, and that the XBB.1.5 monovalent vaccine increases the capacity of serum antibodies to neutralize contemporary variants. IgG, IgA, and total IgG/A/M antibody titers were boosted, which likely includes expansion of a non-neutralizing compartment that mediates disease severity and longer-term protection through Fc effector functions^5^. Indeed, the XBB.1.5 monovalent vaccine was reported to reduce risk of COVID-19 hospitalization by 76.1% in Denmark^6^. In the United States, analysis of two CDC vaccine effectiveness (VE) data networks estimated 52% (95% CI: 47-57%) and 43% (27-56%) VE against hospitalization^7^. This study therefore supports public health recommendations to stay up to date with adapted COVID-19 vaccines.

## Data Availability

All data produced in the present study are available upon reasonable request to the authors.

## Acknowledgements

The authors thank the many participants in this study for their generous contribution. We also gratefully acknowledge the efforts of the entire OHSU COVID-19 serology study team. This work was funded in part by NIH R01AI141549 (to FGT).

---

**Supplemental Figure 1.**
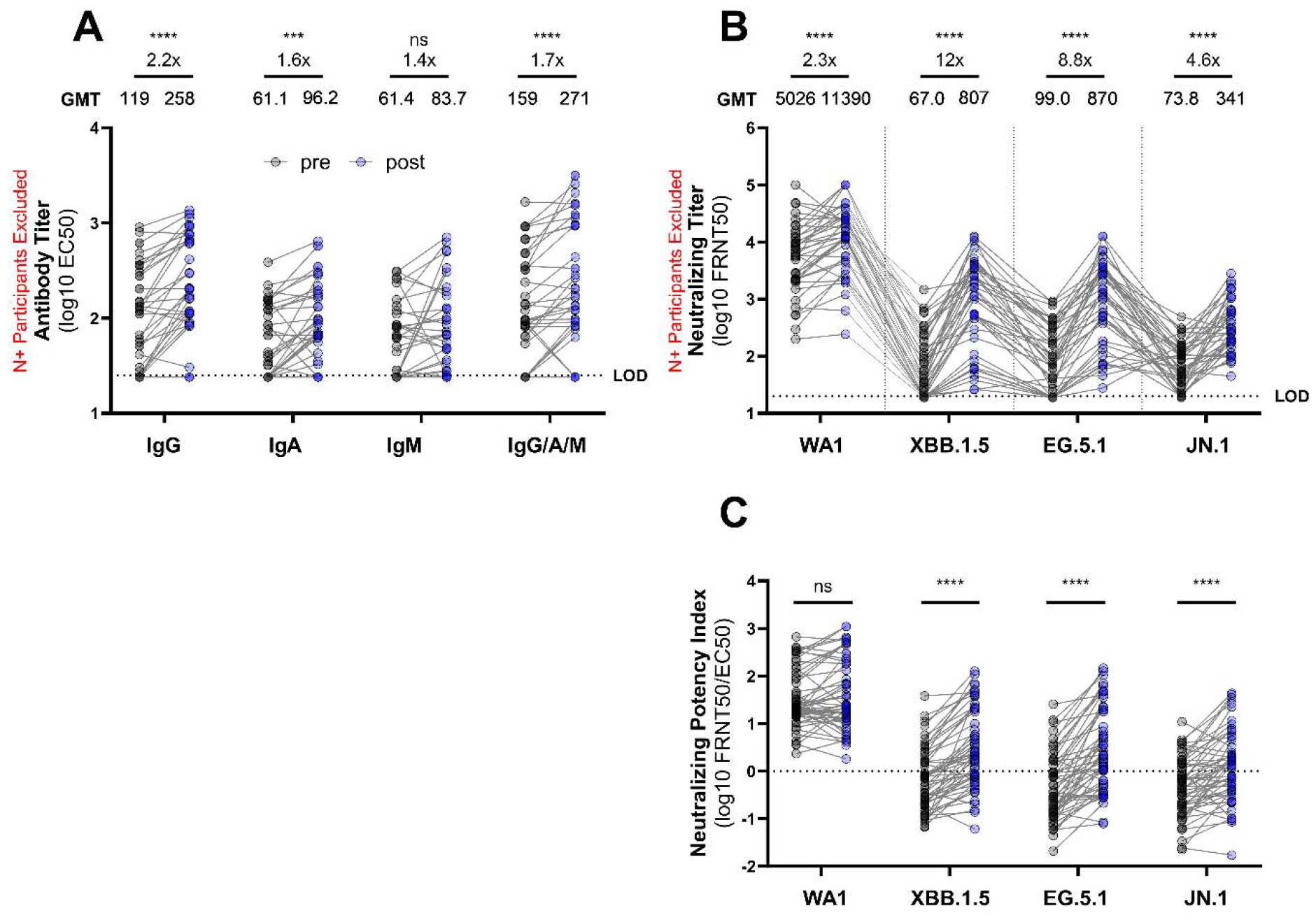
XBB.1.5 monovalent vaccination boosts antibodies in anti-nucleocapsid seronegative participants and neutralization potency index (NPI) against contemporary variants in the overall cohort. Participants with likely prior or current infection (positive for anti-nucleocapsid antibodies) were excluded from analysis in (a-b). (a) Serum antibody isotype titers against spike RBD were determined by ELISA and reported as EC50. (b) Live SARS-CoV-2 neutralization by serum antibodies was assessed by FRNT and reported as FRNT50s. The dotted lines indicate assay lower limits of detection. Geometric mean titers (GMT) for each bar were calculated in GraphPad Prism. Boost ratios were calculated by dividing the post-XBB.1.5 vaccination GMT (post) by pre-vaccination GMT (pre). (c) NPIs for pre-vaccination (pre) and post-vaccination (post) serum samples were calculated by dividing the FRNT50 against the live SARS-CoV-2 variant by total IgG/A/M EC50 for a given participant. The dotted line represents NPI = 1. Reported p-values are the result of restricted effect maximum likelihood models (a,c) or one-way repeated measures ANOVA (b) with Šídák’s multiple comparisons tests (****, p<0.0001; ***, p<0.001; ns=not significant, p>0.05). All individual data points are displayed as filled circles.

## Notes

The authors have declared that no conflicts of interest exist.

### Competing Interest Statement

The authors have declared no competing interest.

### Funding Statement

This study was funded in part by NIH R01AI141549 (to FGT).

### Author Declarations

The institutional review board of Oregon Health & Science University gave ethical approval for this work.

