## Supplemental Methods for "Humoral Immunity Elicited by the XBB.1.5 Monovalent COVID-19 Vaccine"

### Supplemental Materials and Methods

#### Serum collection

At enrollment, 4-6mL of whole blood were collected from each participant, centrifuged at 1000 x g, heat-inactivated at 56°C for 30 minutes, and stored at -20°C as a baseline (pre-boost) serum sample. Approximately 21 days after administration of the XBB.1.5-adapted monovalent vaccine (Moderna Therapeutics), whole blood was collected and similarly processed as a post-boost serum sample.

#### SARS-CoV-2 growth and titration

SARS-CoV-2 clinical isolates were obtained from BEI Resources: [WA1] USAWA1/2020 (NR-52281); [XBB.1.5] USA/MD-HP40900/2022 (NR-59104); [EG.5.1] USA/MD-HP47946/2023 (NR-59503); [JN.1] USA/New York/PV96109/2023 (NR- 59693). Passage 1 virus were generated by inoculating ~70% confluent Vero E6 cells with clinical isolates and cultured until cytopathic effect was observed (72 hours for WA1, 96 hours for XBB.1.5, EG.5.1, JN.1). Culture supernatants were collected and centrifuged at 1000 x g for 10 minutes before freezing in aliquots at -80°C. Passage 2 virus were similarly generated using passage 1 virus to inoculate. Titers were determined by focus-forming assay using 10-fold dilutions. Confluent 96-well plates of Vero E6 were treated with virus serially diluted in dilution media (OptiMEM supplemented with 2% FBS, 1% NEAA, 1%P/S) and incubated at 1 hour at 37°C, 5% CO2. Following infection, overlay media (dilution media with 1% methylcellulose) was added to each well and the plate was incubated for 24 hours for WA1 or 48 hours for XBB.1.5, EG.5.1, and JN.1. Overlay media were then removed and cells were fixed for 1 hour in 4% formaldehyde in PBS. To develop foci, cells were permeabilized in perm buffer (0.1% BSA, 0.1% saponin in PBS) for 30 minutes at RT and incubated with 1:5000 (WA1) or 1:500 (XBB.1.5, EG.5.1, and JN.1) polyclonal anti-SARS-CoV-2 alpaca serum overnight at 4°C. Plates were washed three times with wash buffer (0.01% Tween-20 in PBS) and incubated with 1:20,000 (WA1) or 1:10,000 (XBB.1.5, EG.5.1, and JN.1) anti-alpaca IgG HRP (Novus #NB7242) for 2 hours at RT. Plates were washed three times with wash buffer and 50ul of KPL TrueBlue substrate (Seracare #5510-0030) were added to each well. Plates were incubated for 20 minutes at RT and imaged with a CTL Immunospot Analyzer. Foci were then counted with the Viridot package for R 4.3.1[1].

#### ELISAs

ELISAs for each Ig isotype were performed in biological duplicate in 96-well plates (Maxisorp #423501). Plates were coated with 100uL/well with purified wild-type SARS-CoV-2 RBD at 1ug/mL in PBS and incubated overnight at 4C, rocking. Plates were then washed three times with wash buffer (0.05% Tween-20 in PBS) and blocked with 150ul/well blocking buffer (5% nonfat dry milk powder and 0.05% Tween-20 in PBS) at RT for 1 hour, rocking. Participant sera were diluted in blocking buffer to produce 5 x 4-fold dilutions (1:25 to 1:6,400). After incubating at RT for 1 hour, rocking, plates were washed three times. HRP conjugated to goat anti-human IgG (BD Biosciences #555788), IgA (BioLegend #411002), or IgM (Bethyl Laboratories #A80-100P) were diluted in blocking buffer at 1:3,000 and applied at 100ul/well. Alternatively, HRP conjugated to goat anti-human IgG/A/M (Invitrogen, #A18847) was diluted at 1:10,000 to detect total antibodies. Plates were incubated at RT for 1 hour, rocking, and washed three times prior to adding o-phenylenediamine dihydrochloride (ThermoFisher Scientific, #34005) according to manufacturer instructions. The reaction was stopped after 25 minutes using an equivalent volume of 1M HCl. Optical density (OD) was measured at 492nm using a CLARIOstar plate reader. OD492 reads were normalized by subtracting the average of the negative control wells. For detection of anti-nucleocapsid antibodies, the same protocol outlined above was followed with the exception of coating plates with 1ug/mL nucleocapsid protein (SinoBio #40588-V08B), followed by detection with the HRP goat anti-human IgG/A/M secondary. Serum samples were considered positive for anti-nucleocapsid antibodies if the lowest dilution (1:25) produced an OD492 measurement that was greater than 4 times the average of negative control wells.

**SARS-CoV-2 FRNT**

Focus reduction neutralization tests were performed as previously described[2]. Briefly, participant serum samples were diluted in duplicate using dilution media to make 5 x 10-fold dilutions for WA1 (1:10 to 1:100,000) or 5 x 5-fold dilutions for XBB.1.5, EG.5.1, and JN.1 (1:10 to 1:6250). Equal volumes of diluted serum and 2X diluted virus were combined and incubated for 1 hour at RT. Virus-serum mixes were then added to confluent Vero E6 cells in 96-well plates and incubated for 1 hour at 37°C, 5% CO2 before adding overlay media and incubating for 24 hours (WA1) or 48 hours (XBB.1.5 and EG.5.1). Cells were fixed with 4% formaldehyde in PBS for 1 hour at RT. Staining, developing, imaging, and counting were carried out as for the titration experiment above. For the FRNT, a target of ~30 focus-forming units (FFU) of virus per well is incubated with serum. For every serum dilution series replicate (5 test wells), a $6^{th}$ control well is included that does not contain serum. The number of foci in each test well is divided by the average number of foci in all control wells on one plate to produce a percent neutralization value for each test well.

**ELISA EC50 and FRNT50 calculations**

ELISA OD492 measurements or FRNT percent neutralization values were processed in python (v3.7.6) with numpy (v1.18.1), scipy (v1.4.1), Matplotlib (v3.1.3), and pandas (v1.0.1) data analysis libraries to calculate ELISA EC50 and FRNT50. Replicate data were consolidated and fit with a three-parameter logistic model. For ELISA EC50 and FRNT50 values, replicate curves were generated separately for quality control. Final ELISA EC50 values below the lower limit of quantification of 25 were set to 24 while values above the upper limit of 6,400 were set to 6,401. Final FRNT50 values below the lower limit of quantification of 20 were set to 19 while values above the upper limit of quantification of 200,000 (WA1) or 12,500 (XBB.1.5 and EG.5.1) were set to 200,001 and 12,501, respectively.

**Statistical analysis**

For antibody titers (EC50) and neutralizing potency indices, a restricted effect maximum likelihood (REML) model with Šídák's multiple comparisons tests were performed on the log-transformed data. The REML is analogous to a repeated measures one-way ANOVA, but allows for missing datapoints in repeated measures. For neutralizing antibody titers, a repeated measures one-way ANOVA followed by Šídák's multiple comparisons tests were performed on log-transformed FRNT50 data. Pairwise comparisons were drawn between pre- and post-vaccination titers for each Ig isotype and variant to assess boosting.
